# LUKB: Preparing Local UK Biobank Data for Analysis

**DOI:** 10.1101/2024.05.05.24306906

**Authors:** Xiangnan Li, Shuming Wang, Hui Zhang, Zixin Hu

## Abstract

**Background:** While UK Biobank data holds immense potential for human health research, its complex pre-processing steps involving decryption, extraction, and code mapping often act as a barrier for researchers, diverting them from their core research questions. A freely available tool for preparing UK Biobank data would reduce the workload of researchers and the costs produced by alternatively using UK Biobank Research Analysis Platform (RAP).

**Results:** We developed LUKB, an R Shiny-based web tool that simplifies UK Biobank data preparation by automating pre-processing tasks. Through simple actions, researchers can add downloaded UK Biobank data to LUKB, achieving rapid data decryption, efficient extraction, and accurate code mapping effortlessly.

**Conclusion:** LUKB reduces pre-processing time, allowing researchers to dedicate more time to their scientific endeavors, and provide an alternative to UK Biobank RAP to minimize costs. LUKB is freely available at Github (https://github.com/HaiGenBuShang/LUKB).

## Background

UK Biobank is a treasure trove of medical data, encompassing genetics, proteomics, imaging, and medical records for over 500,000 participants [1-4]. This wealth of information has facilitated countless medical breakthroughs [5-8] and continues to fuel new discoveries. As scientific inquiry into human health deepens, the utilization of UK Biobank data becomes increasingly pivotal, a trend underscored by the growing number of research endeavors centered around this invaluable resource (see **Supplementary Figure S1, Additional file 1**).

However, navigating the vast data within UK Biobank can be challenging, posing obstacles to research speed and efficiency. Processing downloaded encrypted UK Biobank data often involves complex steps (**Figure 1a**), potentially requiring multiple data applications and reprocessing if the data doesn’t meet expectations. Moreover, data previews are unavailable before data processing, which might lead to frustration when downloaded data proves unsuitable. While the UK Biobank Research Analysis Platform (RAP) allows data previewing, downloading the actual data incurs costs, potentially restricting research scope.

**Figure 1.**
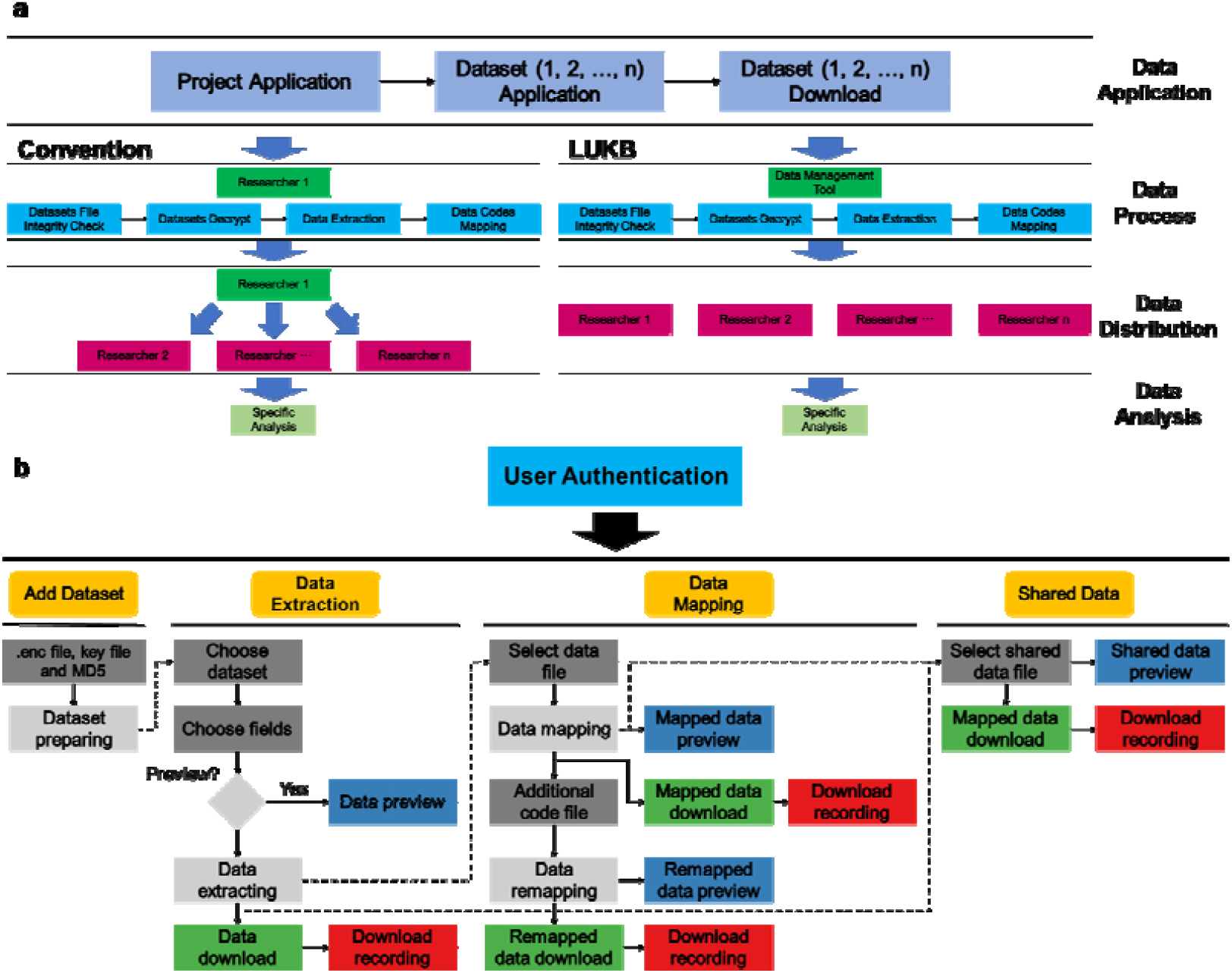
LUKB facilitates UK Biobank data preparation. a) Steps for UK Biobank data analysis. The bottom-left side shows the conventional steps, while the bottom-right side illustrates the steps using LUKB. b) LUKB data processing steps and design.

Here we provide LUKB, a freely deployable R Shiny-based web tool, which empowers researchers to overcome these hurdles and maximize the potential of UK Biobank data. Our locally deployed solution seamlessly integrates data access, previewing, and sharing within research groups with secure and offline data access. LUKB simplifies the workflow by:

- Decrypting and unpacking encrypted UK Biobank data.
- Extracting interested data from decrypted files.
- Previewing the interested data, saving valuable time and effort.
- Mapping extracted data to human-readable formats.
- Sharing and collaborating with colleagues within research groups.

By eliminating potentially unnecessary application steps and providing instant data previews, LUKB paves the way for faster, more efficient research while minimizing costs. This user-friendly tool empowers both seasoned and novice researchers to unlock the full potential of UK Biobank data and contribute to life-changing discoveries.

## Implementation

### Tool Design

The intuitive design of LUKB empowers researchers to access UK Biobank data with five key components (**Figure 1b**). **User Authentication** safeguards data by filtering access, upholding ethical research practices. **Add Dataset** effortlessly integrates downloaded data, streamlining workflows. **Data Extraction** lets researchers focus on their research by pinpointing specific fields of interest. **Data Mapping** transforms raw data into their real-world meaning by deciphering cryptic codes and providing descriptive titles. Finally, **Shared Data** fosters collaboration by securely storing files for exchange between colleagues, accelerating scientific breakthroughs.

### User Authentication

LUKB prioritizes responsible data management by ensuring only authorized researchers access UK Biobank data. A simple text file securely stores usernames and passwords, allowing data holders to easily manage access. This secure setup safeguards sensitive information while remaining user-friendly for adding or removing researchers.

### Add Dataset

This component handles encrypted UK Biobank data (.enc files) with ease, seamlessly decrypting them using corresponding key files (.key) and verifying their integrity through MD5 string checks. The decrypted data is then passed to the **Data Extraction** component for further analysis. Simultaneously, a comprehensive dictionary is created, containing field descriptions, item counts, and data-coding indices to guide researchers through the dataset’s structure.

### Data Extraction

After passing through the **Add Dataset** component, the decrypted data can be previewed and extracted. Researchers could provide the fields of interest and preview the first 100 records or extract the data directly. To optimize performance and prevent tool unresponsiveness, the data extraction tasks are submitted to the operating system, circumventing R’s single-task limitation. Once the extraction task completion signal is detected, the extracted data is passed to the **Data Mapping** component for download. To further protect data, download recording is triggered for all download actions, aiding in the identification of any abnormal downloads.

### Data Mapping

The extracted data from some fields is less readable for humans as it consists solely of codes and numbers. To enhance readability, it’s essential to transform these codes into their real-world meanings. UK Biobank provides multiple code mapping files to assist in this process. This component leverages the most comprehensive code mapping file to perform the transformation, providing instant previews of the mapped data. However, in cases where certain fields remain unmapped, researchers could upload additional coding files for those specific fields, ensuring a comprehensive mapping of all available data.

### Shared Data

Each downloadable data file can be shared and found within this component. When choosing to share data files, a short comment about the file is required to facilitate other researchers’ understanding of its contents.

## Results and discussion

### LUKB simplifies the preparation of analysis-ready data from local UK Biobank data

Traditionally, accessing and preparing UK Biobank data for analysis is a complex maze of applications, downloads, and manual processing. LUKB streamlines this journey, transforming local UK Biobank data into analysis-ready data with just a few clicks. Previously, the UK Biobank data would be downloaded to local computers after a successful data application. Researchers would then carry complicated data processing steps using computer commands, including data file integrity checks, decryption, specific data extraction, and code mapping if necessary (**Figure 1a**). The subsequent data sharing often involved hard drives or USB flash drives. This process, repeated for each new data application and data updates, were both tedious and time-consuming. With LUKB, researchers can bypass time-consuming data preparation and dive straight into unlocking life-changing discoveries (**Figure 1b**).

### Limitations

UK Biobank offers two main data types: rectangular (spreadsheet-like) and bulk/genomic (e.g., images, raw sequencing data). While valuable, bulk/genomic data require specialized tools to analyze due to their unstructured formats. Therefore, LUKB focuses on the majority of rectangular data, ensuring a familiar format that researchers can readily use for their analyses.

### Recommendations

Since LUKB can manage data of multiple fields, we recommend that researchers who use this tool apply for and import as much data as possible to streamline workflows and minimize data approval processes. We also recommend deploying this tool on the intranet to avoid malicious internet access and enhance data protection.

## Conclusion

Our R Shiny-based tool, LUKB, empowers researchers by streamlining the preparation of UK Biobank data, facilitating efficient analyses, and potentially leading to groundbreaking research advancements. LUKB simplifies the intricate process of data preparation, including decryption, extraction, and code mapping, allowing researchers to dedicate more time to their scientific inquiries. For detailed information on deployment and usage, please refer to the supplementary materials (**Additional file 1**).

## Data Availability

All data produced in the present work are contained in the manuscript

## Availability and requirements

Project name: LUKB

Project home page: https://github.com/HaiGenBuShang/LUKB

Operating system(s): Linux

Programming language: R, Bash

Other requirements: Internet browser

License: MIT

Any restrictions to use by non-academics: None

## List of abbreviations

RAP: Research Analysis Platform

## Declarations

### Ethics approval and consent to participate

Not applicable.

### Consent for publication

Not applicable.

### Availability of data and materials

Project name: LUKB

Project home page: https://github.com/HaiGenBuShang/LUKB

Operating system(s): Linux

Programming language: R, Bash

Other requirements: Internet browser

License: MIT

Any restrictions to use by non-academics: None

### Competing interests

The authors declare that they have no competing interests.

### Funding

This work was supported by the Shanghai Rising-Star Program (21QB1400900), and the National Natural Science Foundation of China (32100510) and has been conducted using the UK Biobank Resource under Application Number 103791.

### Author contributions

X.L. conceived the idea, designed and developed the application, and drafted the manuscript. H.Z. supervised the project. S.W. and H.Z. contributed to the development of the application. All authors read and approved the final manuscript.

## Acknowledgements

The authors thank all the authors who contributed to the R shiny or related packages and the funding agencies that supported this project.

## Notes

### Competing Interest Statement

The authors have declared no competing interest.

### Funding Statement

This study was funded by the Shanghai Rising-Star Program (21QB1400900), and the National Natural Science Foundation of China (32100510) and has been conducted using the UK Biobank Resource under Application Number 103791.

