## Supplementary Data for "LUKB: Preparing Local UK Biobank Data for Analysis"

**Supplementary Figure**

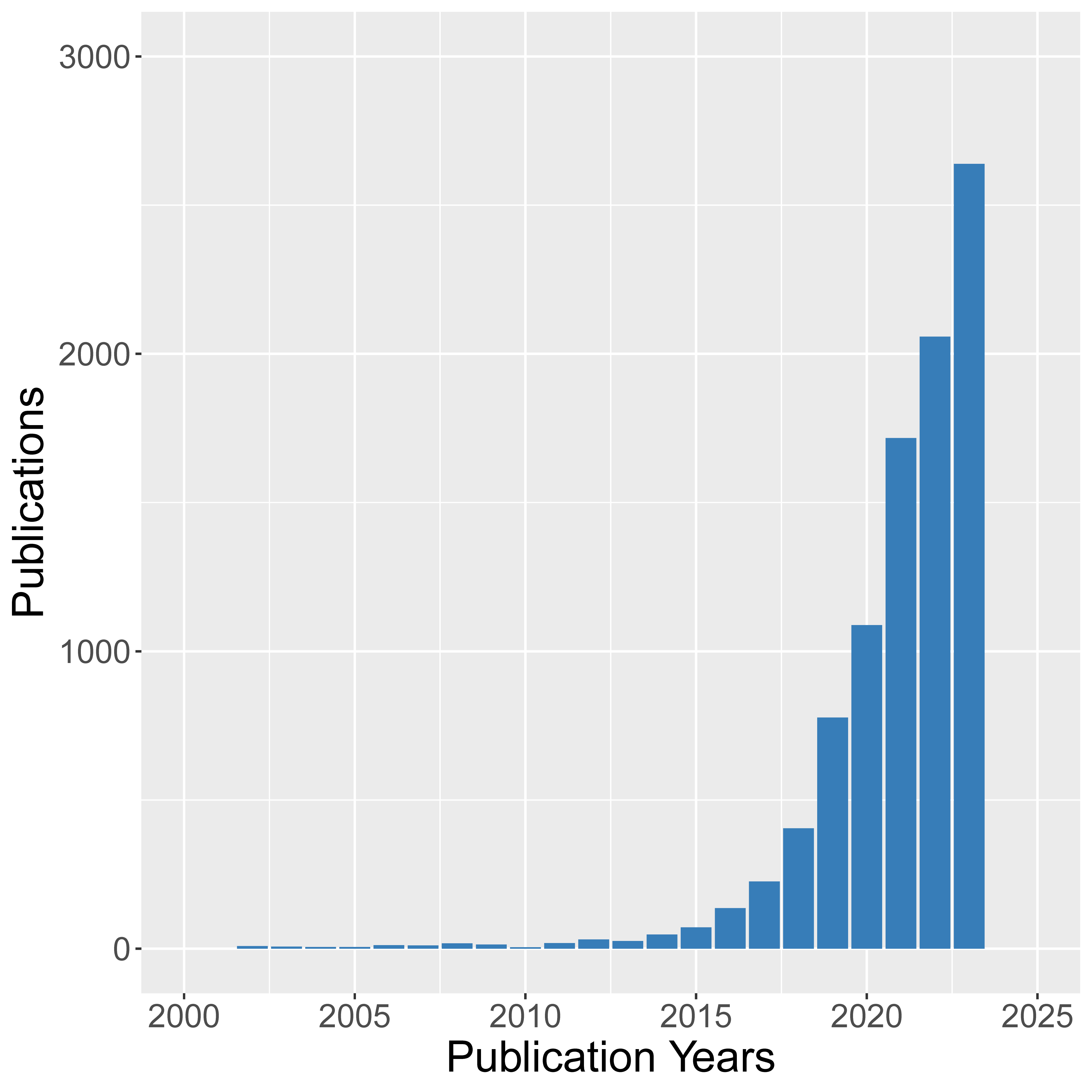

**Figure S1. Publications related to “UK Biobank” are rapidly increasing recent years.** Figure S1 shows the publications indexed by Web of Science under the topic of “UK Biobank”, reflecting the rapid increase in research involving this valuable resource.

**Supplementary materials**

**LUKB Deployment and usage**

0. **Dependencies**

LUBK was developed using R version 4.2.3, so it is recommended to install R 4.2.3 or higher. Some required packages can be installed by executing the following command at the main directory:

*Rscript required_packages.R*

1. **Add Users and Start LUKB**

LUBK will start automatically after user information is added. LUKB uses a simple text file to store user information. After downloading the source code, change to the main directory, and then execute:

*chmod +x add_users.sh && ./add_users.sh*

Then type the user name, password, and TRUE or FALSE to set data file download permission (Figure S2). LUKB will start in a short time. Researchers can access LUKB via http://your_server_ip:1111/.

The default port used by LUKB is 1111. Make sure this port is added to your firewall rules. To add port 1111, execute:

*sudo iptables -I INPUT -p tcp --dport 1111 -j ACCEPT (Ubuntu)*

or

*sudo firewall-cmd --add-port=1111/tcp (CentOS)*

Then port 1111 will be opened temporarily. To change the default port, change “port = 1111” to “port = the_port_you_want” at the bottom line in the app.R file.

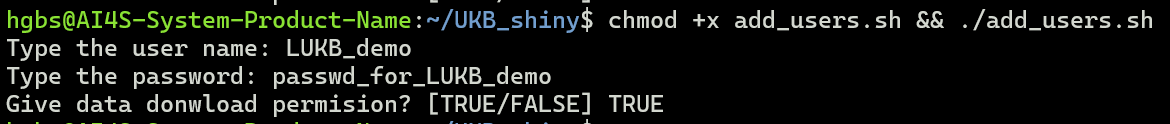

**Figure S2. Add user information.**

2. **Access to LUBK**

As a web-based tool, LUKB is easily accessible. To access LUKB, open the link http://your_server_ip:1111/ in the web browser (**Figure S3**). Once authenticated, researchers can access data through the user interface.

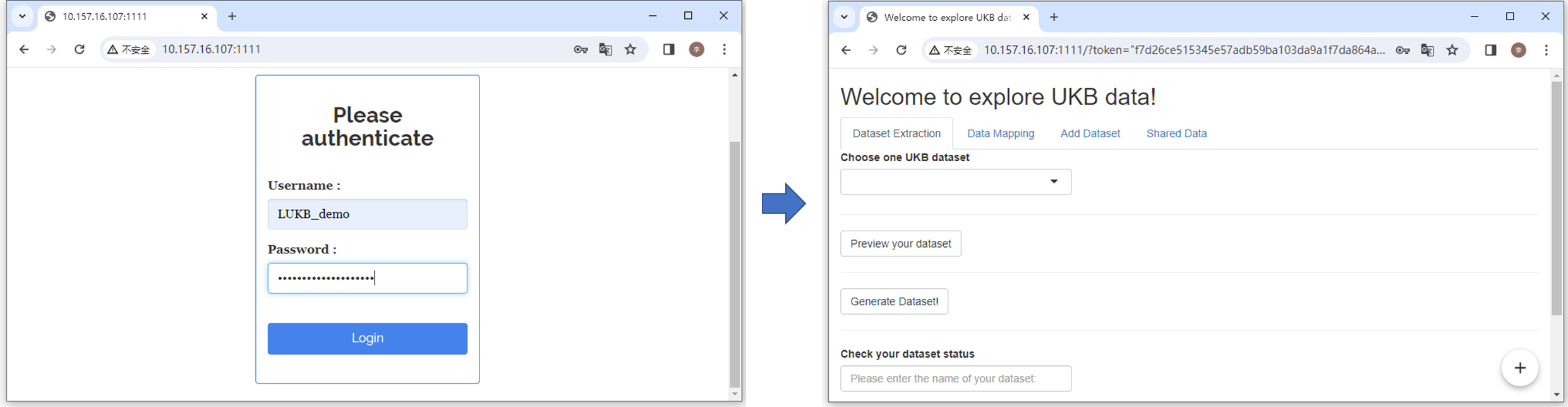

**Figure S3. Access to LUKB.** By open the link (*http://your_server_ip:1111/*) in the web browser, researchers can access to LUKB.

3. **Add Dataset**

For initial use, researchers must import UK Biobank data into LUKB. Typically, UK Biobank data is downloaded in a file named ukbrun_id.enc, where run_id corresponds to your specific data basket. This file is accompanied by a key file and an MD5 checksum. To add data to LUKB, researchers should upload the downloaded .enc file, the corresponding key file, and provide the MD5 string. Once these are entered, the data will be imported into LUKB (**Figure S4**).

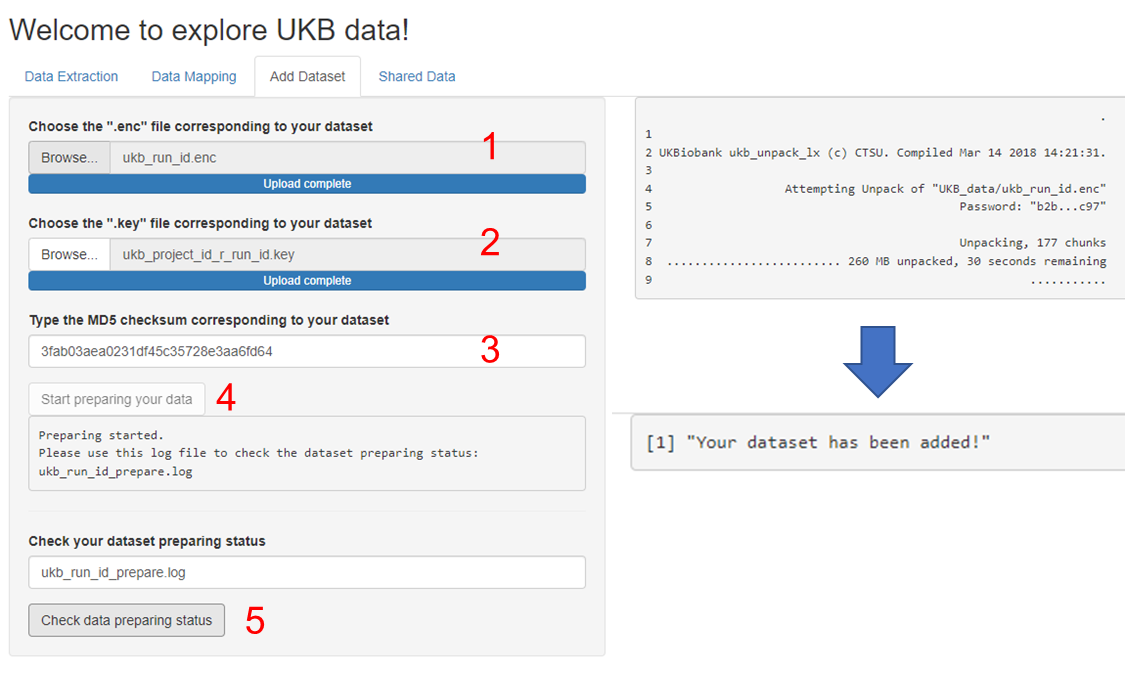

**Figure S4. Add dataset to LUKB.** Researchers can add UK Biobank datasets to LUKB by providing the .enc file, key file, and MD5. The right panel displays the data preparation status upon clicking the “Check data preparation status” button. The message “Your dataset has been added!” indicates successful dataset addition to LUKB. 1) upload .enc file. 2) upload key file. 3) provide MD5 string. 4) submit dataset adding task to operating system. 5) check dataset adding task status.

4. **Data Extraction**

Once data importation is complete, researchers can extract interested data by selecting specific fields or providing field IDs. LUKB also allows researchers to preview the data of these selected fields before extraction. For large datasets, users can monitor the extraction status by clicking the "Check extraction status" button. After data extraction is complete, researchers with data download permission can download the extracted data (**Figure S5**).

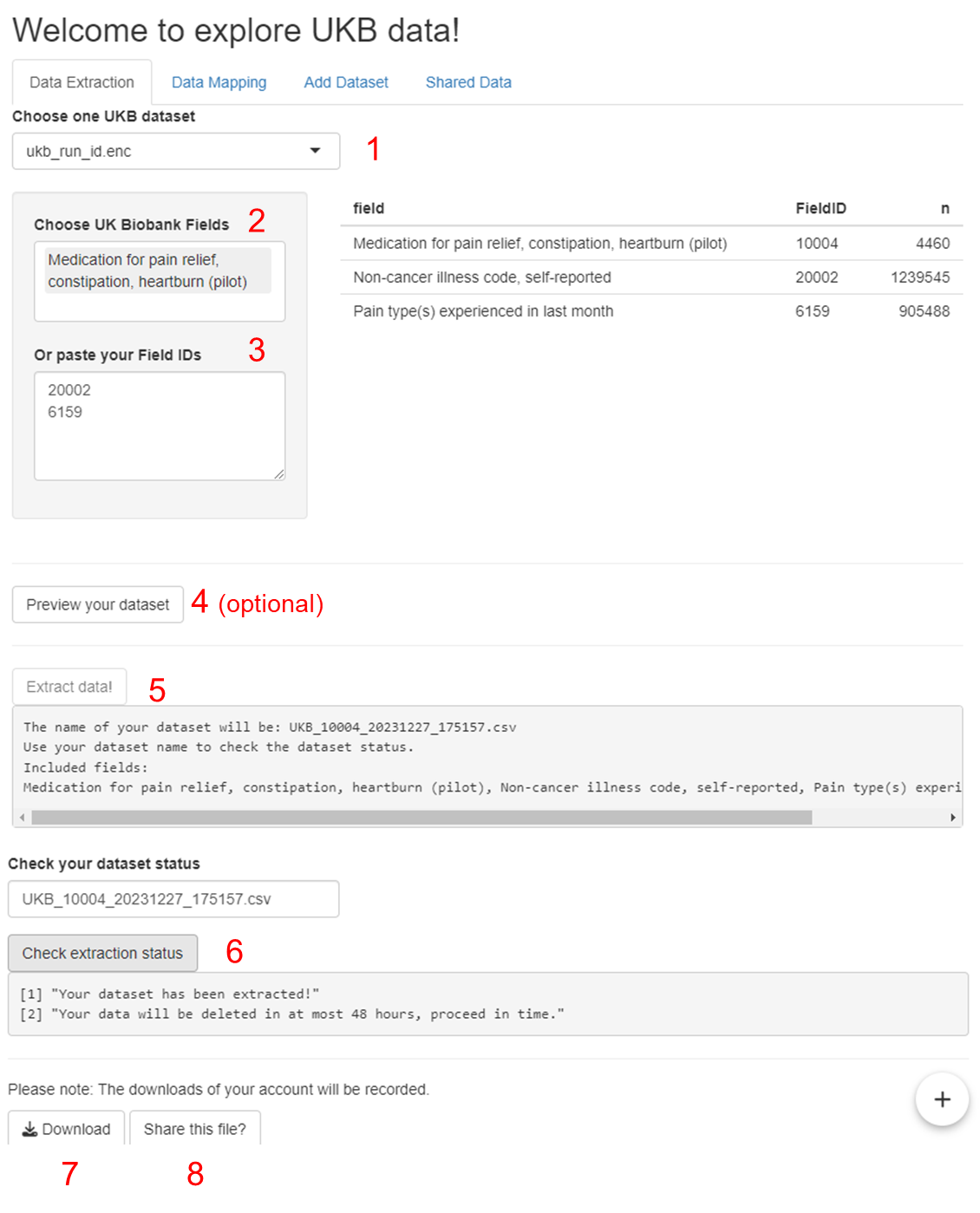

**Figure S5. Extracting Data from an Imported Dataset.** After selecting a dataset, researchers can either choose specific fields or provide field IDs to extract the interested data. 1) choose the dataset for extraction. 2, 3) provide fields or field IDs for data extraction. 4) preview the data of provided fields (optional). 5) submit data extraction task to operating system. 6) check data extraction task status. 7) download the extracted data. 8) choose to share the extracted data file.

5. **Data Mapping**

In some cases, extracted data may not be directly readable due to the use of codes, such as in the field “Pain type(s) experienced in last month” (Field ID: 6159). For example, “College or University degree” might be represented as “1” in this field. To enhance readability, LUKB allows researchers to map these codes to their real meanings. To perform code mapping, click the "Data Mapping" button after uploading or choosing a data file. After mapping, researchers can preview the remapped data. However, some fields might still be unmappable. This typically occurs when the mapping information is stored in separate coding files. For instance, the coding information for “Non-cancer illness code, self-reported” (Field ID: 20002) is stored in the Data-Coding 6 file. To map these fields, researchers need to upload the relevant coding files and specify the field to be mapped (**Figure S6**).

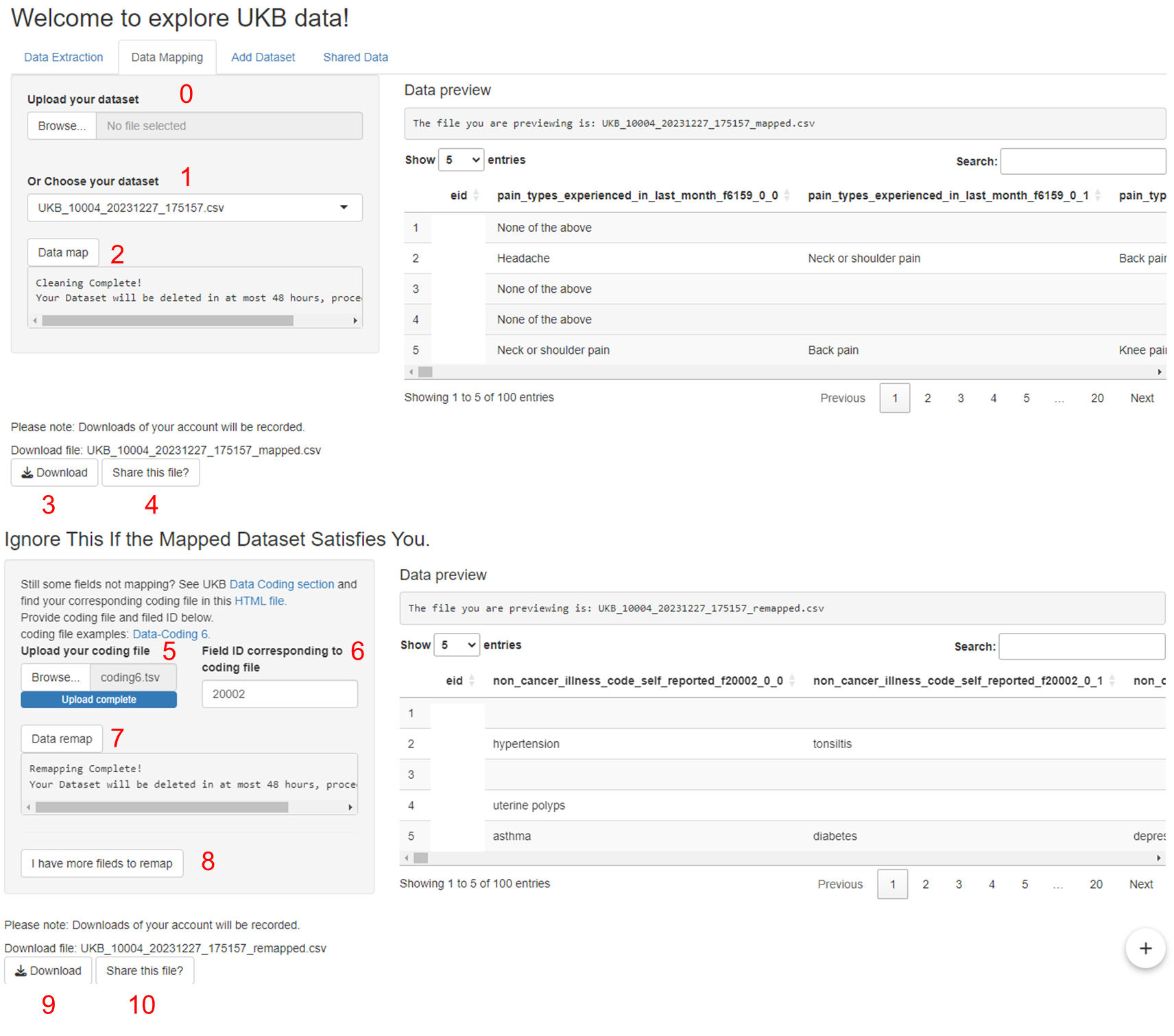

**Figure S6. Mapping codes to their real meanings.** 0) upload data file produced in Data Extraction component. 1) choose the file for code mapping. 2) submit code mapping task. 3) download the mapped data file. 4) choose to share the mapped data file. 5) upload relevant data coding file. 6) specify the field ID corresponding to the coding file. 7) submit code remapping task. 8) optionally, add more coding file and fields to map, this step can be repeated multiple times. 9) download the remapped data file. 10) choose to share the remapped data file.

6. **Data Sharing**

LUKB encourages researchers to share extracted or mapped data with others, as unshared files are automatically removed after a maximum of 48 hours to conserve system storage. Upon completing data extraction or mapping, researchers can share a file by clicking the “Share this file?” button. Adding optional remarks about the file’s contents can help other researchers quickly familiarize themselves with the shared data. All shared files can be viewed and downloaded within the “Shared Data” component (**Figure S7**).

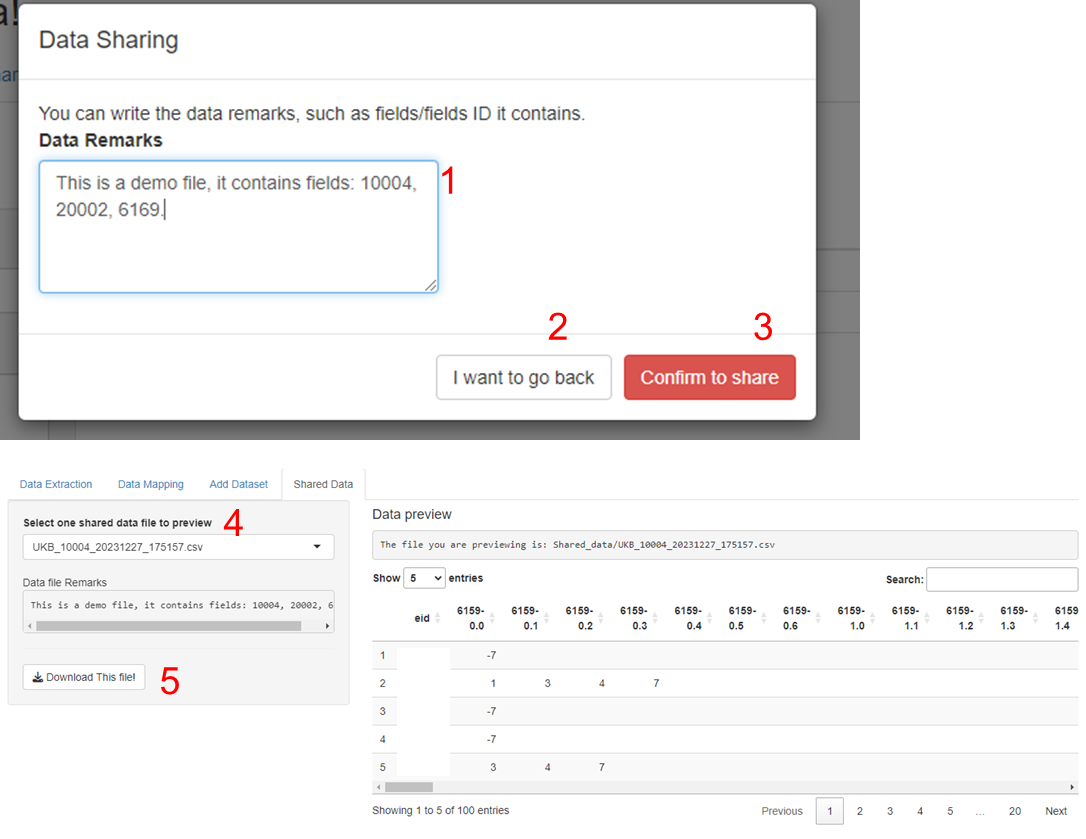

**Figure S7. Shared data file remarks and download.** Each shared data file is accompanied by a descriptive file providing context and insights. When choosing to share a file, researchers should provide a descriptive overview. 1) description information. 2) cancel data file sharing. 3) confirm data file sharing. 4) choose one shared data file. 5) download the shared data file.

7. **Downloading Monitor**

LUKB maintains comprehensive records of user downloading activities for tracking and auditing purposes. These records, including downloaded files, specific fields extracted, and corresponding user information, can be found within the “Logs/” directory.
